# Global Cancer Burden Attributable to Dietary Risks: Trends, Regional Disparities, and Future Projections (1990-2050)

**DOI:** 10.1101/2024.11.30.24318246

**Authors:** Jinghao Liang, Yijian Lin, Zishan Huang, Jingchun Ni, Hongmiao Lin, Yiwen Cai, Jihao Qi, Liangyi Yao, Luoyao Yang, Dianhan Lin, Zhihua Guo, Weiqiang Yin, Jianxing He

**Author notes:** These authors contributed to this work equally (Jinghao Liang, Yijian Lin, Zishan Huang). Corresponding authors: Weiqiang Yin; Jianxing He, Department of Thoracic Surgery and Oncology, the First Affiliated Hospital of Guangzhou Medical University, State Key Laboratory of Respiratory Disease & National Clinical Research Center for Respiratory Disease, Guangzhou, 510120, China.

## Abstract

Cancer remains a leading global cause of death, with its burden increasingly shaped by demographic shifts and dietary factors. This study utilized the Global Burden of Disease (GBD) 2021 database to evaluate cancer burdens attributable to dietary risks from 1990 to 2021, accounting for differences by age, gender, region, and socioeconomic level, and projected trends through 2050 using a Bayesian Age-Period-Cohort model. The global disability-adjusted life years (DALYs) attributable to dietary risks declined from 302.48 to 189.62 per 100,000 population (AAPC: −1.49%) over three decades, yet disparities remain prominent across Socio-Demographic Index (SDI) regions. High-SDI countries, such as Luxembourg, achieved substantial reductions, while low-SDI nations like Lesotho and Zimbabwe experienced rising burdens, driven by inadequate dietary quality and limited health resources. Key dietary risks, including low intake of whole grains, milk, and red meat, demonstrated improvement in high-income countries but worsening trends in many low- and middle-income regions. Projections suggest a continued global decline in cancer burden attributable to dietary factors by 2050, with high-income regions benefiting most, while Latin America, the Caribbean, North Africa, and the Middle East may experience slower progress or transient increases. Additionally, the burden of poor dietary practices is expected to rise sharply among individuals aged 75 years and older, underscoring the compounding effects of aging populations. These findings highlight the urgent need for culturally tailored dietary interventions and evidence-based policies to address disparities, reduce cancer burdens, and improve outcomes for vulnerable populations globally.

## Introduction

Cancer remains a significant global public health issue and is the second leading cause of death worldwide. Between 1990 and 2019, age-standardized incidence and mortality rates for cancer demonstrated a decreasing trend. However, the onset of the COVID-19 pandemic led to a subsequent increase in global age-standardized mortality rates in 2020 and 2021, partially reversing previous progress^1^. The International Agency for Research on Cancer (IARC) projects that the number of new cancer cases worldwide will rise from approximately 20 million in 2022 to over 35 million by 2050—an increase of 77%^2^, underscoring the immense disease burden faced by the global population.Understanding modifiable risk factors for cancer, particularly those related to dietary habits that are relatively easier to intervene upon, is essential for informing cancer prevention and control strategies. Diet has been established as a major modifiable risk factor for cancer in multiple studies^3^. Adherence to the Mediterranean diet, characterized by an emphasis on vegetables, fruits, whole grains, nuts, seeds, legumes, moderate consumption of fish, olive oil, and alcohol, and reduced intake of red or processed meats and dairy products, has been shown to reduce cancer risk^4–6^. Nevertheless, prior research has often focused on single dietary patterns or nutrients, and largely centered on digestive system cancers such as esophageal and colorectal cancer^7–10^. There remains a lack of comprehensive analysis evaluating various dietary factors across all cancer types and geographic regions, particularly regarding the impact of dietary disparities across different regions on cancer burden.

To address this research gap, this study evaluates cancer risk attributable to dietary factors from 1990 to 2021, incorporating potential confounding factors such as gender, age, region, and socioeconomic level. Leveraging the Global Burden of Disease (GBD) database, this study provides a comprehensive assessment of the impact of dietary factors on the global cancer burden, encompassing a wide range of cancer types beyond the digestive system. Using a Bayesian Age-Period-Cohort (BAPC) model, we aim to project the impact of dietary risk factors on cancer incidence and mortality trends globally and regionally through 2050. Our findings will inform evidence-based dietary adjustments in different regions to reduce cancer risk and mortality, providing actionable recommendations to mitigate the future cancer burden associated with diet.

## Methods

### Data source

The present study utilized the latest data from the GBD 2021 database, a comprehensive global health repository encompassing detailed information on 371 diseases, 88 risk factors, and numerous injuries^11,12^. The primary data sources for GBD 2021 include vital registration systems, verbal autopsies, surveys, censuses, surveillance systems, and cancer registries, providing critical evidence for estimating disease incidence and mortality rates.

### Definition

We identified 9 dietary risk factors (detailed in the supplementary table s1) that satisfied the GBD selection standards for inclusion as risk factors. These criteria involve the significance of the risk factor in terms of disease burden or policy impact, the availability of adequate data to estimate exposure, the strength of epidemiological evidence supporting a causal association between exposure and health outcomes, and the availability of data to quantify the magnitude of this association per unit change in exposure. Additionally, evidence must support the generalizability of these effects across different populations. The process for evaluating the epidemiological evidence of causality for each diet-disease pair is comprehensively documented elsewhere and summarized in the appendix^11^.

### Global cancer burden estimates

Colon and rectum cancer, stomach cancer, breast cancer, tracheal, bronchus, and lung cancer, and esophageal cancer data, including neoplasm-related deaths, DALYs, and corresponding age-standardized rates, were obtained from the Global Health Data Exchange (GHDx) website (https://vizhub.healthdata.org/gbd-results/). The DALYs and mortality for these neoplasms were classified using the International Classification of Diseases, Tenth Revision (ICD-10)^13^(supplementary table s2). Prostate cancer data, which predominantly consisted of negative values, was excluded from the analysis due to its limited interpretability.

### Statistics

In this study, Joinpoint regression analysis was performed using Joinpoint 5.1.0 to compute the annual percentage change (APC) and average annual percentage change (AAPC) in cancer mortality and DALYs rates^14^. This widely applied statistical model facilitates the identification of significant turning points in disease trends, as well as overall patterns over specified time intervals. Decomposition analysis was employed to quantify the individual contributions of population age structure, population growth, and epidemiological changes to cancer-related disability-adjusted life years (DALYs) associated with dietary risk, providing a clear understanding of these factors’ influence on the overall cancer burden^14^. Additionally, Pearson’s correlation coefficient was calculated to assess the relationship between the Socio-Demographic Index (SDI) and age-standardized cancer DALYs^15,16^. To evaluate cross-country health inequalities, we used the slope index of inequality and the concentration index to measure both absolute and relative health disparities. The slope index was derived by regressing cancer incidence, mortality, and DALYs on a relative social position scale based on GDP per capita, with heteroskedasticity controlled using a weighted regression model. The concentration index was calculated by fitting the observed cumulative distribution of the population by income to the Lorenz curve for cancer burden, followed by numerical integration of the area under the curve^17,18^. Finally, the Integrated Nested Laplace Approximation (INLA) framework combined with the Bayesian Age-Period-Cohort (BAPC) model was used to predict future trends in cancer burden. The BAPC model, based on Global Burden of Disease (GBD) data from 1990 to 2021 and population projections from the World Health Organization, provides accurate forecasts while addressing convergence issues common to traditional Bayesian Markov Chain Monte Carlo (MCMC) methods^19^. All statistical analyses and data visualizations were performed using R 4.4.1, with statistical significance defined at P < 0.05.

## Result

### Global Reduction and Regional Disparities in Diet-Related Cancer Burden

Based on the Global Burden of Disease (GBD) database, the global disability-adjusted life years (DALYs) attributable to dietary risk factors for cancer decreased substantially from 302.48 per 100,000 population in 1990 to 189.62 per 100,000 in 2021, with an average annual percentage change (AAPC) of −1.49% (95% CI: −1.57 to −1.42). This trend indicates a significant reduction in the global burden of diet-related cancers over the past three decades. Notably, Kazakhstan (DALY AAPC: −3.25%), China (−2.57%), Turkmenistan (−2.81%), and Luxembourg (−2.30%) demonstrated the greatest reductions in cancer burden. Conversely, the burden increased in Lesotho (+2.21%), Zimbabwe (+1.08%) and Romania (+0.80%). At the Socio-Demographic Index (SDI) regional level, countries in high-SDI regions exhibited a marked declining trend in DALYs (−1.50%), including Austria (−2.34%) and Luxembourg (−2.30%). Middle-SDI regions demonstrated greater heterogeneity, with substantial improvements in Kyrgyzstan (−2.53%) and Uzbekistan (−2.38%) but a rising burden in countries such as the Philippines (+0.70%) and Romania (+0.80%). Low-SDI regions displayed similarly diverse trends, with significant reductions in Burundi (−1.57%) and Rwanda (−1.55%) but marked increases in Zimbabwe and Lesotho, underscoring disparities in health resource allocation and intervention intensity (table1).

### Global Trends in Cancer Burden Attributable to Top Three Dietary Risk

Globally, the leading dietary risk factors contributing to the cancer burden were diet high in red meat, diet low in milk, and diet low in whole grains, each demonstrating considerable geographic and temporal variability. The burden of diet high in red meat decreased significantly in high-income regions, particularly in Europe and North America countries, with AAPCs between −1.48% and −2.31%, while a contrasting increasing trend was noted in low- and middle-income regions, including sub-Saharan Africa, South America, and Southeast Asia, with AAPCs up to 2.34% (Fig. 1 A-C, supplementary table s3). The cancer burden attributable to Diet low in milk showed significant declines in North America, Europe, and Oceania (AAPC ranging from −2.05% to −19.2%), whereas positive AAPCs were observed in South Asia, Africa, and the Caribbean, indicating insufficient dairy consumption in these regions(Fig. 1 D-F, supplementary table s3). Low whole grain intake demonstrated a declining burden in high-income regions such as North America and Australia (AAPC from −2.32% to −1.40%), while increasing trends persisted in Latin America, the Middle East, and sub-Saharan Africa (AAPC between 0.60% and 2.70%) (Fig. 1 G-I, supplementary table s3).

**Fig. 1.**
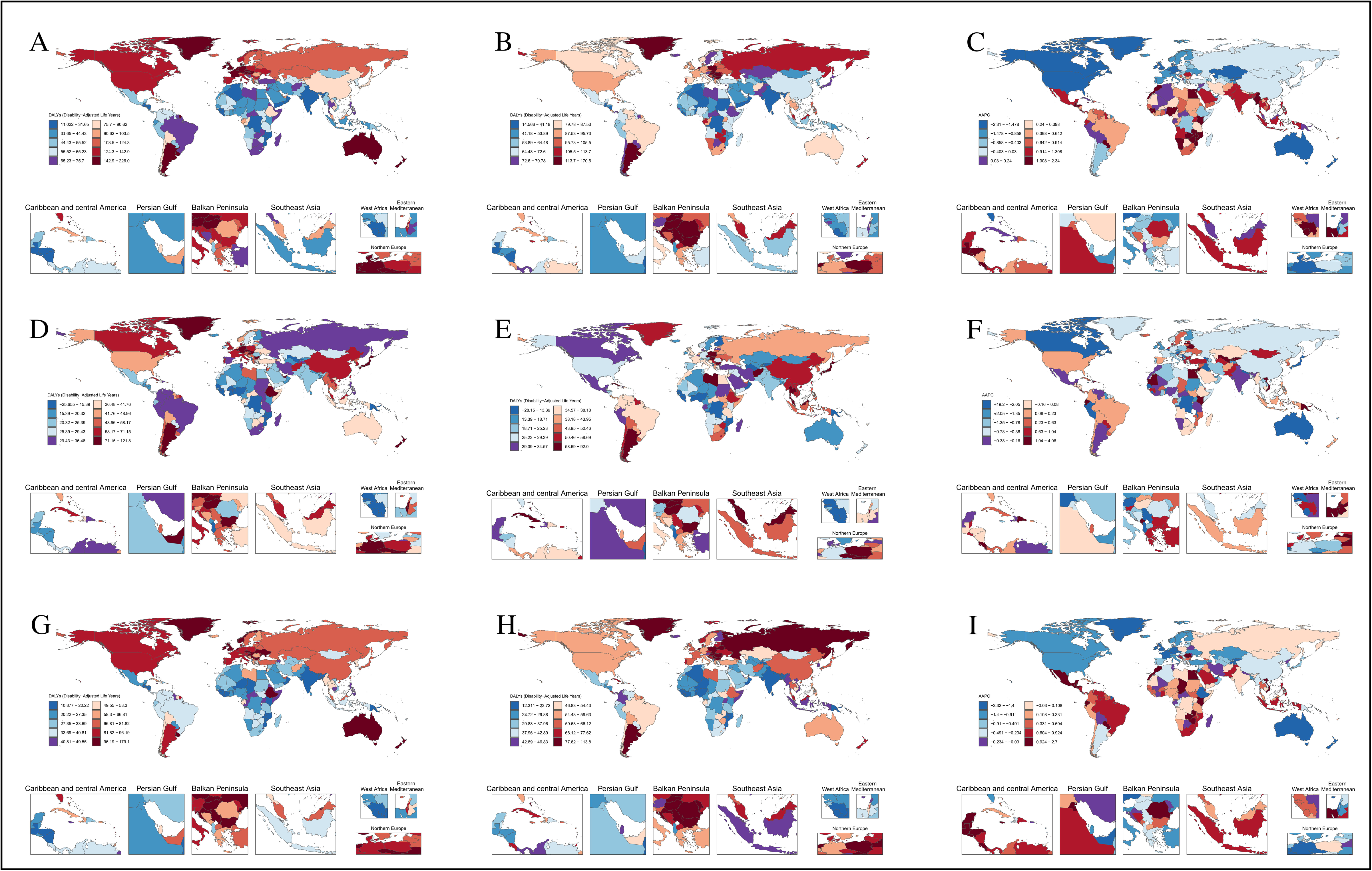
Comparison of cancer DALYs attributable to the three most common dietary risks across 204 countries and territories worldwide. (A-C) Cancer DALYs attributable to a diet high in red meat for the years 1990, 2021, and the average annual percentage change (AAPC) from 1990 to 2021. (D-F) Cancer DALYs attributable to a diet low in milk for the years 1990, 2021, and the AAPC from 1990 to 2021. (G-I) Cancer DALYs attributable to a diet low in whole grains for the years 1990, 2021, and the AAPC from 1990 to 2021.DALYs: Disability-adjusted life years; AAPC: Average annual percentage change.

### Specific Dietary Risk Factors and Their Associations with Cancer Types

In 2021, poor dietary practices remained major contributors to cancer-related DALYs globally. For instance, high red meat consumption was associated with a breast cancer burden of 28.37 DALYs (95% CI: −0.0092 to 60.54). Colorectal cancer showed a significant burden attributable to several dietary factors:Diet low in whole grains (50.19 DALYs; 95% CI: 20.37-76.30), diet low in fiber (3.58 DALYs; 95% CI: 1.58-5.50), diet high in processed meat (15.11 DALYs; 95% CI: −3.60 to 30.93), diet low in calcium (24.70 DALYs; 95% CI: 18.17-31.02), and diet low in milk (42.99 DALYs; 95% CI: 11.73-71.23). Additionally, gastric cancer was linked to diet low in vegetables (20.78 DALYs; 95% CI: −4.68 to 102.38) and diet high in sodium (44.53 DALYs; 95% CI: −7.45 to 222.31), while diet low in fruits was linked to the burden of tracheal, bronchial, and lung cancer (18.46 DALYs; 95% CI: 9.49-26.90).(supplementary table s4) There were no significant associations observed between the cancer burden and other dietary factors, such as diet high in trans fatty acids, diet low in omega-6polyunsaturated fatty acids, diet low in seafood omega-3 fatty acids, diet low in legumes,diet low in nuts and seeds, or diet high in sugar-sweetened beverages.

### Socioeconomic Disparities(SDI) and Shifting Patterns in Diet-Related Cancer Burden

The global diet-related cancer burden is predominantly driven by colorectal cancer, particularly in high-SDI regions, while gastric and esophageal cancers contribute significantly in low-SDI regions. This pattern highlights the interplay between dietary habits and levels of socioeconomic development, with the cancer burden shifting towards colorectal and breast cancers as SDI increases (Fig. 2 A-B, supplementary table s7). The inter- and intra-regional disparities in dietary risk-related cancer burden across 204 countries and 21 regions further underscore the role of socioeconomic context. Low-SDI regions showed relatively stable intraregional variation but significant interregional differences, whereas middle- and high-SDI regions demonstrated substantial variability both within and across regions. Central Asia exhibited particularly pronounced intraregional disparities, with DALY rates ranging from approximately 200 to over 400 per 100,000 population (Fig. 2 C-D, supplementary table s6). From 1990 to 2021, the association between SDI and DALY rates for diet-related cancers exhibited an increasing trend, with the slope of the relationship rising from 272.16 (95% CI: ∼222.18-322.15) in 1990 to 299.17 (95% CI: ∼258.70-339.63) in 2021. The concentration index (CI) for diet-related cancer burden remained negative in both years, at −0.17 in 1990 and −0.18 in 2021, indicating better health outcomes among disadvantaged populations. However, the absolute increases in both slope and CI values indicate that inequality in cancer burden attributable to dietary risk factors has worsened over time (Fig. 2 E-F, supplementary table s7).

**Fig. 2.**
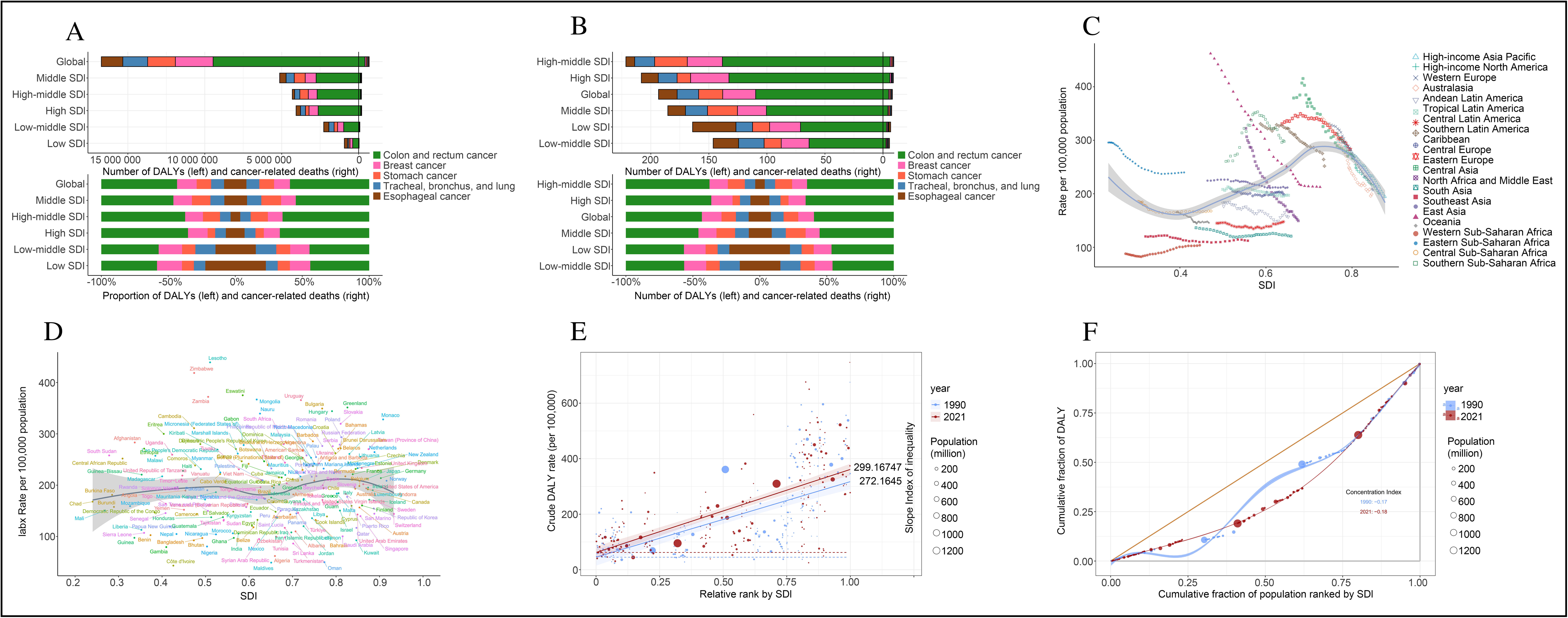
Association between cancer and the Socio-demographic Index (SDI). (A) Global and region-specific cancer cases for all ages, and (B) age-standardized rates, alongside the proportion of DALYs and deaths by cancer type. (C) Association between age-standardized DALYs rate and SDI across 21 regions, and (D) association with SDI across 204 countries. (E) Slope indexes and (F) concentration indexes for cancer DALYs globally from 1990 to 2021.SDI: Socio-demographic Index.

### Age, Gender, and Decomposition Analysis of Diet-Related Cancer Burden

Age-stratified analysis indicated that the burden of diet-related cancers increases significantly with age, particularly among individuals aged 75 years and older. In the 75-79 age group, DALYs exceeded 50,000 in both males and females, with cancer-related mortality peaking in this cohort. Males experienced a higher burden across most age groups, highlighting a disproportionate impact of diet-related cancers on men (Fig. 3 A-B, supplementary table s8). Decomposition analysis further revealed that population growth and aging were the primary drivers of the increased cancer burden attributable to dietary factors, while improvements in epidemiological factors partially mitigated the overall impact. Population growth contributed approximately 1 million DALYs, while aging accounted for an additional increase of 500,000 DALYs (Fig. 3 C-D, supplementary table s9).

**Fig. 3.**
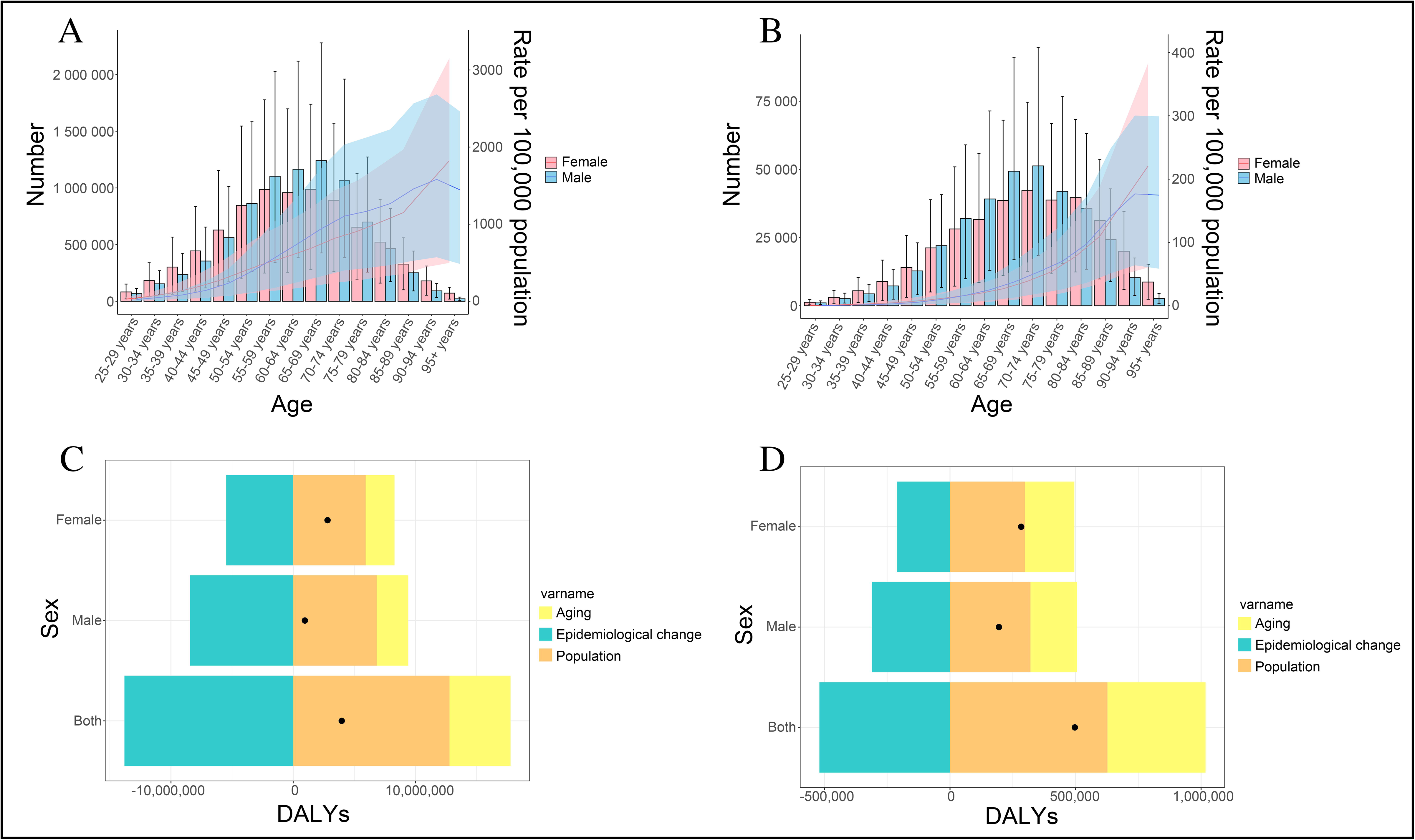
Global burden of cancers by sex. (A) Global age-specific counts and rates of disability-adjusted life years (DALYs) by sex, and (B) global age-specific counts and rates of cancer deaths by sex. (C) Decomposition analysis of trends in cancer DALYs by sex from 1990 to 2021, and (D) decomposition analysis of trends in cancer deaths by sex from 1990 to 2021.

### Projected Trends in Cancer DALYs Attributable to Dietary Risks Up to 2050

Projections for dietary risk-related cancer DALYs from 2022 to 2050 suggest a global decline in age-standardized cancer DALYs, from 344.27 to 223.71 per 100,000 population. High-income regions are projected to exhibit the steepest decline, while a relatively slower rate of decline is anticipated in Latin America and the Caribbean. A transient rebound in cancer mortality is expected in North Africa and the Middle East between 2025 and 2030. While the burden is expected to decrease continuously among individuals aged 25-54, a sharp increase is projected for the elderly population, particularly in the 75-95 age group, reflecting the significant impact of population aging on the future cancer burden attributable to dietary Factors(Fig. 4, supplementary table s10).

**Fig. 4.**
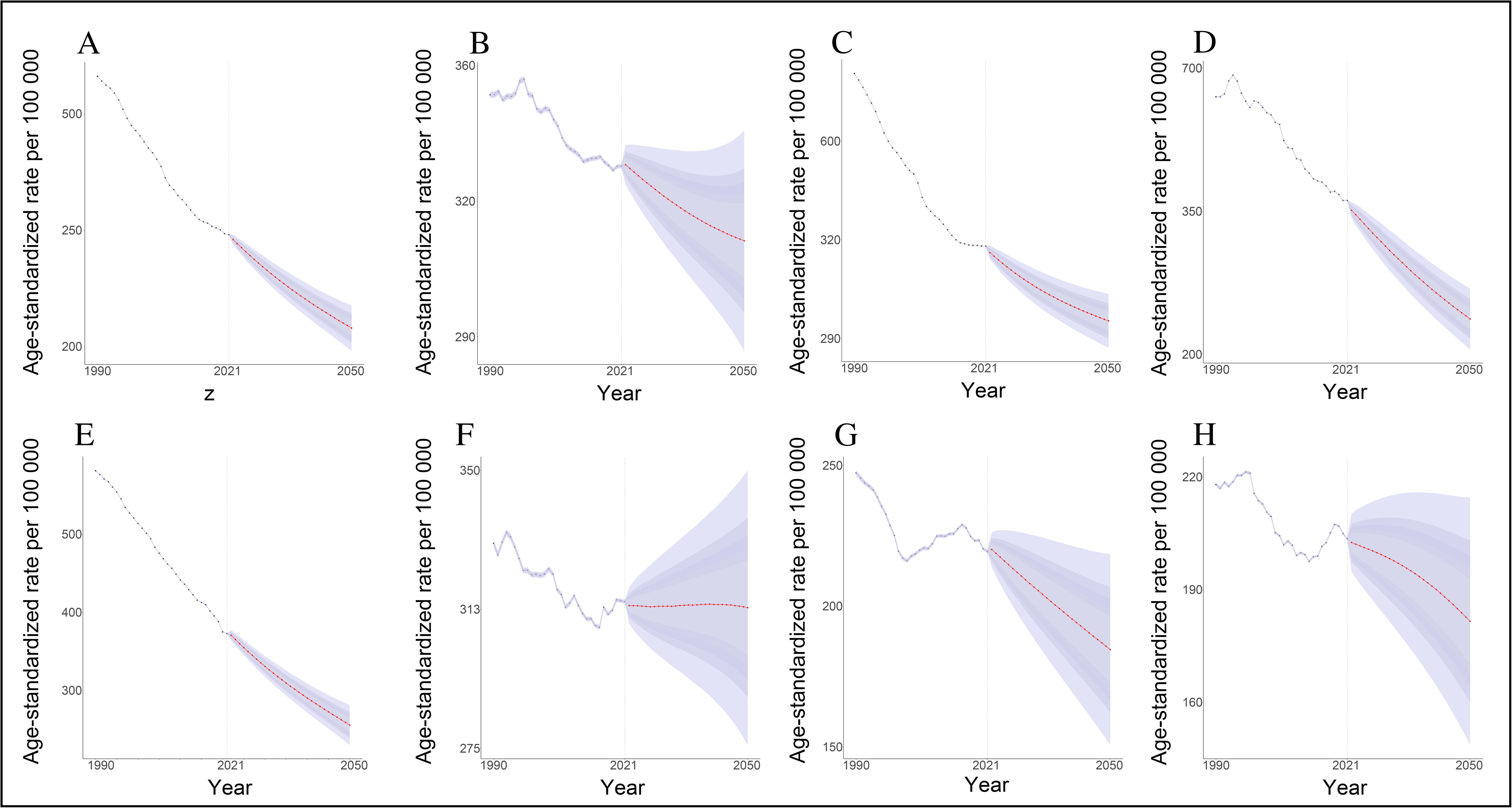
Projections of cancer DALYs by 2050 based on the BAPC model.(A) Projected global cancer DALYs, (B) projections for Sub-Saharan Africa, (C) projections for Southeast Asia, East Asia, and Oceania, (D) projections for Central Europe, Eastern Europe, and Central Asia, (E) projections for high-income countries, (F) projections for Latin America and the Caribbean, (G) projections for North Africa and the Middle East, and (H) projections for South Asia.DALYs: Disability-adjusted life years; BAPC: Bayesian Age-Period-Cohort model.

## Discussion

Our study, based on the Global Burden of Disease (GBD) data from 1990 to 2021, provides an in-depth analysis of the impact of dietary factors on cancer burden and its changing trends, revealing significant regional disparities influenced by socioeconomic and dietary characteristics. Over the past three decades, the global cancer burden attributable to dietary factors has decreased by an average of 1.49% annually. However, the degree of improvement is uneven across different regions and populations. High Socio-Demographic Index (SDI) countries such as Austria and Luxembourg have significantly reduced the burden of colorectal cancer through dietary optimization and screening measures, whereas low-SDI countries like Lesotho and Zimbabwe continue to experience rising burdens of gastric and esophageal cancers due to poor dietary nutrition and insufficient resources. Additionally, the burden of diet-related cancers varies significantly by gender and age, with males and individuals aged 75 years and older being disproportionately affected. These findings underscore the importance of culturally tailored health education programs, dietary optimization, increased intake of key nutrients, and the promotion of dietary-related cancer screening measures as essential pathways to reducing the global cancer burden.

The impact of dietary factors on specific cancers is highly targeted, providing a scientific basis for precision interventions through understanding the complex underlying pathophysiological mechanisms. Low whole grain intake, for example, reduces dietary fiber, disrupts gut microbiota, and increases the risk of exposure to carcinogens such as nitrosamines and bile acids, thereby significantly increasing the incidence of colorectal cancer^20^ ^21^. Low calcium intake weakens the protective function of epithelial cells, heightening the risk of esophageal cancer ^22^ ^23^. In high-income countries, policies promoting whole grain and calcium-enriched foods have yielded substantial success; for instance, Northern Europe has effectively reduced the burden of colorectal cancer ^24^ and esophageal cancer ^25^ through food subsidies and health labeling policies. Conversely, sub-Saharan Africa and Southeast Asia face persistently high burdens of gastric and esophageal cancers due to insufficient calcium and dietary fiber intake ^26^, coupled with high salt and pickled food consumption ^27^ ^28^. Addressing this requires the implementation of regional policies that promote the consumption of calcium-rich and fiber-rich foods, the introduction of legumes and root crops, the development of affordable calcium-fortified foods, and strengthened health education targeting high-salt and pickled food consumption to achieve dietary improvements and reduce disease burden.

Interestingly, our data also reveal that high trans-fatty acid intake, low omega-6 polyunsaturated fatty acid intake, low seafood omega-3 fatty acid intake, and high sugar-sweetened beverage intake do not show a significant association with cancer burden, in contrast to previous research that has highlighted their carcinogenic potential. Prior studies suggest that trans fats may increase ovarian cancer risk by inducing inflammation ^29^, and excessive omega-6 intake may interfere with omega-3 fatty acid utilization, thereby promoting tumorigenesis ^30^ ^31^. These discrepancies may reflect the complexity of cancer etiology, the overall effect of dietary patterns, or the dilution of associations for specific regions or subpopulations in global analyses.

Our findings further indicate that differences in adverse dietary factors across geographic, age, and gender dimensions necessitate more precise strategies for intervention. At the geographic level, high-SDI countries have reduced the burden of diet-related cancers through long-term policy interventions, such as the European “Healthy Food Label Program” ^32^, which has increased the selection of whole grain and low-fat food options among residents. In contrast, low-SDI countries continue to bear a high burden of diet-related cancers, particularly in regions with high incidence rates of gastric and esophageal cancers ^33^, due to a lack of health resources and weak dietary education. For these countries, international aid or regional cooperation could support community-based dietary improvement programs that teach methods for preparing low-cost, nutrient-dense diets ^34^ ^35^ ^36^. Regarding age distribution, the burden of diet-related cancers increases significantly with age, particularly among individuals aged 75 years and older, underscoring the importance of dietary interventions during middle age to prevent cancer burden in later life. Gender differences also show that males bear a higher burden of diet-related cancers compared to females, likely related to dietary behaviors and higher red meat consumption ^37^ ^38^. Thus, policy interventions for male populations should focus on limiting high-risk foods, such as processed meats and high-fat snacks, and promoting healthy alternatives.

The carcinogenic mechanisms of adverse dietary factors differ significantly across SDI regions, underscoring the importance of targeted policy interventions. In high-SDI countries, processed foods, high-fat diets, and low dietary fiber intake are the main risks ^39^ ^40^. These countries can reduce the burden of diet-related cancers by restricting the sale of processed foods, optimizing nutrition standards, and providing subsidies for healthy foods. Northern Europe’s subsidy for whole grain foods, which has significantly increased the prevalence of healthy diets, serves as a successful example ^41^. In low-SDI countries, deficiencies in key nutrients, such as calcium and dietary fiber, are the primary carcinogenic drivers ^42^, closely linked to traditional single dietary patterns and poverty. Promoting the cultivation of fiber-rich crops through agricultural policy and improving calcium intake through school nutrition programs are recommended interventions. Meanwhile, middle-SDI countries face the dual challenge of dietary transition, with both high-salt pickled foods and processed foods posing threats. These countries should adopt a “dual-path dietary policy,” which aims to reduce high-salt food consumption through campaigns similar to the salt reduction initiatives in the Americas and Europe ^43^, while also implementing fiscal controls on ultra-processed foods and guiding residents toward healthy alternatives. Such differentiated intervention strategies can effectively address dietary issues across different SDI regions, thereby reducing the global burden of diet-related cancers.

Future projections of the cancer burden attributable to dietary factors highlight significant regional disparities and complexities among different populations, emphasizing the urgent need for global health interventions. Although the age-standardized DALYs for diet-related cancers are projected to decline globally from 344.267 to 223.713 per 100,000, this improvement is not uniform between high-income and low-income regions. For example, high-income countries are expected to experience a further decline in diet-related cancer burden, benefiting from the long-term promotion of whole grain and dairy product consumption ^44^, whereas Latin America and the Caribbean are likely to see only modest declines, indicating gaps in the coverage of intervention measures. Mortality projections for North Africa and the Middle East suggest a temporary rebound in cancer-related deaths between 2025 and 2030, potentially due to the continuation of traditional high-salt dietary practices. Simultaneously, the burden among those aged 75 years and older is expected to rise substantially, underscoring the impact of aging populations. Precision interventions, such as designing nutrient-fortified dietary plans for the elderly, enhancing screening services, and ensuring that health resources can accommodate the needs of an aging population, are crucial for driving improvements in global health outcomes and ensuring equitable distribution of health resources worldwide.

## Data Availability

All data generated in this study are available upon reasonable request from the authors. All data produced in this study are included in the manuscript. All data generated are accessible online.

https://www.healthdata.org/research-analysis/gbd

## Author Contributions

Study conceptualization: Jianxing He, and Weiqiang Yin. Accessed and verified the underlying data reported in the manuscript: Jinghao Liang and Yijian Lin. Data curation: Zishan Huang, Yijian Lin, Jingchun Ni, Jihao Qi, and Hongmiao Lin. Formal analysis: Yiwen Cai, Yijian Lin, Liangyi Yao, and Jihao Qi. Designed Fig.s and tables: Yuanqing Liu, Weijie Yang, and Zishan Huang. Writing original draft of the manuscript: Jinghao Liang, Dianhan Lin, and Yijian Lin. Review and editing of the manuscript: Jinghao Liang, Yijian Lin, Luoyao Yang, Jihao Qi, Jingchun Ni, Yiwen Cai, and Liangyi Yao. Jinghao Liang, Yijian Lin, and Zishan Huang contributed equally to this work. All authors read and approved the final version of the manuscript, had full access to all the data, and are responsible for the decision to submit for publication.

## Declaration of interests

The authors declare no competing interests.

## Data Availability

Data used in this analysis are accessible through the Global Health Data Exchange (GHDx) platform. This study utilizes primary data from the Global Burden of Disease (GBD) 2021, which are available for download online.

## Acknowledgment

This study received no funding.

## Ethics approval and consent to participate

Ethical approval was not required for this study, as it utilized publicly available, anonymized data aggregated at the population level.

## Consent for publication

Not applicable.

## Notes

### Competing Interest Statement

The authors have declared no competing interest.

### Funding Statement

This study did not receive any funding.

